# Examining comorbid and transdiagnostic depression clinical outcomes across anxiety, autism, attention deficit hyperactivity disorder (ADHD), bipolar disorder, depression, and schizotypal personality groups: a novel NeuroMark SPECT approach

**DOI:** 10.64898/2026.04.15.26350953

**Authors:** Amritha Harikumar, Bradley Baker, Daniel Amen, David Keator, Vince D. Calhoun

## Abstract

Major depressive disorder (MDD) is a highly prevalent neuropsychiatric disorder characterized by depressed mood, feelings of sadness, loss of interest, and reduced pleasure related to daily activities. The clinical etiology of depression has been extensively studied, with research indicating biological, social, and psychological factors related to onset of depressive symptoms. Despite increased knowledge related to MDD, there is no tangible biomarker developed for MDD.

Neuroimaging modalities such as single photon emission computed tomography (SPECT) have been utilized to characterize regional cerebral perfusion (rCBF). Functional dysconnectivity in depressed patients have been examined, with depressed individuals showing elevated depression scores and decreased rCBF in cognition and executive functioning networks. While SPECT can be utilized to monitor rCBF changes with respect to symptom severity, it alone cannot be utilized to develop a potent biomarker. Advanced multivariate methods such as independent component analysis (ICA) have been used to visualize disconnected functional patterns across disorders including depression and schizophrenia. Given no current SPECT studies examine transdiagnostic clinical profiles, the current study aims to bridge this gap.

We utilized the 68 NeuroMark SPECT template across six patient groups. Factor scores investigating three key symptoms of depression: worry/rumination, moodiness, and social disinterest, and measured the loading parameter strength (i.e. component expression for each NeuroMark domain/subdomain) across the 68 components were examined. We identified significant relationships between symptoms and frontal, triple network, sensorimotor, and visual components across the three symptom profiles. Future studies should examine these trends across larger sample sizes, and increased clinical samples.

## INTRODUCTION

Major depressive disorder (MDD) is a highly prevalent neuropsychiatric disorder characterized by depressed mood, feelings of sadness, loss of interest, and reduced pleasure related to daily activities (American Psychiatric Association, 2013). The clinical etiology of depression has been extensively studied, with research indicating biological, social, and psychological factors related to onset of depressive symptoms (Drevets, 1998; Z. Li et al., 2021; Zallar & Dupont, 2026). Despite increased knowledge related to MDD, there is no tangible biomarker developed for MDD. Neuroimaging modalities such as single photon emission computed tomography (SPECT) have been utilized to track changes in regional cerebral perfusion or blood flow (rCBF; Goozée et al., 2014; Koyama et al., 1997; J. Li et al., 2018). Functional dysconnectivity in depressed patients has been examined, with depressed individuals showing elevated depression scores and decreased rCBF in cognition and executive functioning networks (J. Li et al., 2018).

While SPECT can be utilized to monitor rCBF changes with respect to symptom severity, it alone cannot be utilized to develop a potent biomarker. Advanced multivariate methods such as independent component analysis (ICA) have been used to visualize disconnected functional patterns across disorders including depression and schizophrenia. Depression SPECT studies have been conducted, however there are currently no transdiagnostic depression studies utilizing SPECT imaging to compare depressive symptoms across other psychiatric disorders. Combining SPECT with ICA methods have proven to be useful in identifying disconnected networks in schizophrenia, including auditory, subcortical, and cerebello-thalamocortical networks (Harikumar et al., 2025). The current study builds from these existing studies to utilize a newly developed NeuroMark SPECT template with NeuroMark 2.2 labels (Jensen et al., 2024).

The goals of the current study are to understand patterns of dysconnectivity in depression vs. healthy controls, specifically across anxiety, bipolar disorder, depression, schizotypal personality, and attention deficit hyperactivity disorder profiles. Previous multimodal fusion studies have identified links between clinical symptoms (e.g. disrupted executive function) and reward processing across schizophrenia, depression, autism, and attention deficit hyperactivity disorder (Qi et al., 2020a; Schwarz et al., 2020). These links have seldomly been explored in SPECT studies. Therefore, the current study seeks to examine relationships between clinical symptomatology and functional dysconnectivity broadly between patients vs. controls, and evidence of attention deficits/anxiety/executive dysfunction across most of the patient groups.

## METHODS

### SPECT Imaging Procedure and Preprocessing

76 healthy controls and a subsample of 2,743 depressed patients with both SPECT images and clinical data were acquired from an initial pool of 22,809 subjects from the Amen Clinic (https://www.amenclinics.com/; see Figures 1-3 and Tables 1a-1b for demographic information). Out of the depression sample, six patient subgroups were selected based on shared symptomatology along with other shared profile characteristics (Ardesch et al., 2023; Barkus & Badcock, 2019; Boisvert et al., 2024; Merola et al., 2024; Qi et al., 2020b; Schwarz et al., 2020).

**Figure 1.**
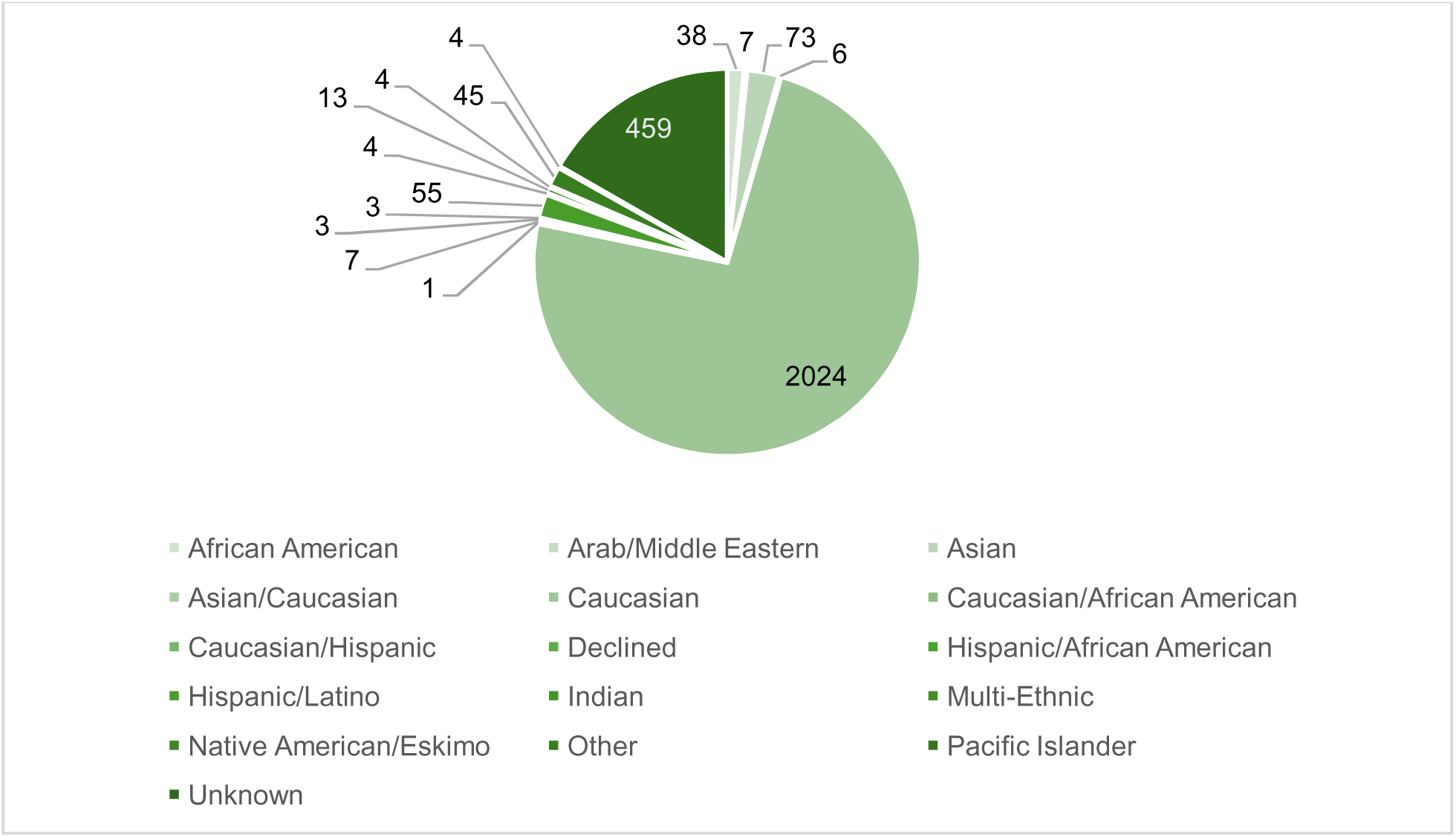
Demographic information for 2,746 patients from the Amen Clinic Dataset, depicted as a detailed pie chart. 3 subjects were classified as biracial or unknown.

**Figures 2 and 3.**
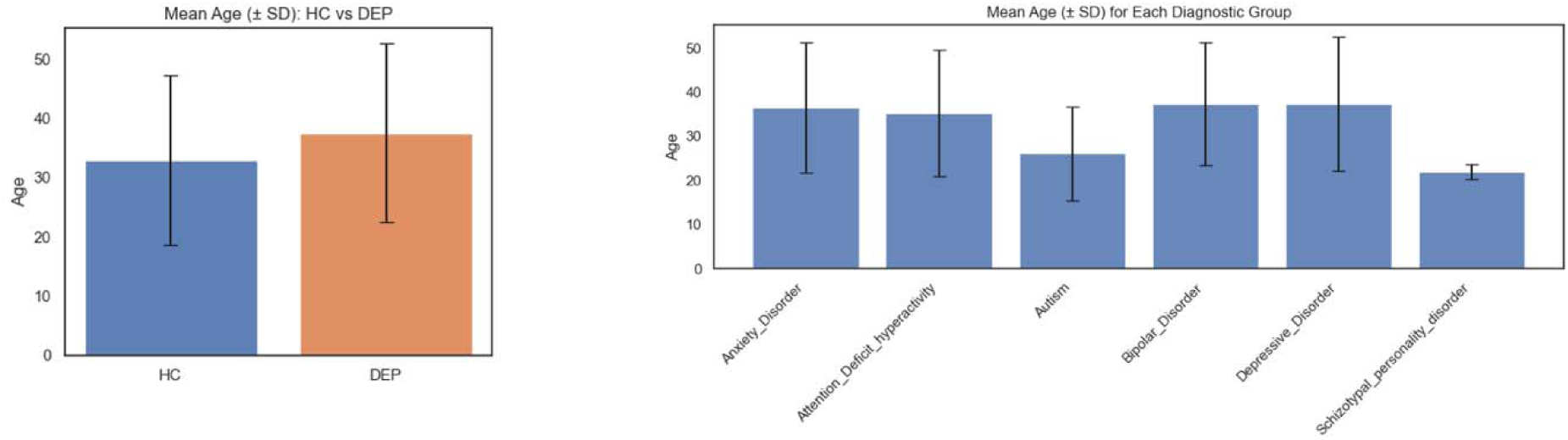
Mean age differences between healthy controls and depressed subjects, with health controls being slightly younger than the depressed cohort. Across the transdiagnostic populations, autism cohorts and schizotypal personality patients skewed the youngest, with anxiety, ADHD, bipolar, and depressed participants ranging similarly across average participant ages.

**Table 1a.** Clinical Information for Patients. Demographic information for patients from the Amen Clinic Dataset, including age, sex breakdown, and information regarding the six disorders of focus for the paper.

| N | Mean Age (SD) | Female % | Male % | Anxiety Disorder | ADHD | Autism | Bipolar Disorder | Depressive Disorder | Schizotypal Personality Disorder |
| --- | --- | --- | --- | --- | --- | --- | --- | --- | --- |
| 2743 | 37.42 (15.068) | 56.61 | 43.38 | 2226 (81.2%) | 1185 (43.2%) | 62 (2.3%) | 173 (6.3%) | 2633 (96.0%) | 3 (0.1%) |

**Table 1b.** Clinical Information of Controls and Racial Breakdowns. Demographic information for healthy controls from the Amen Clinic dataset, with the site breakdown, average age and standard deviation, along with race classification information across sites. 12 sites were included in this study. Healthy controls were all from Site 2.

| Amen Clinics Site # | Age (Mean \| SD) | Subject Breakdown | Gender | Categorization |
| --- | --- | --- | --- | --- |
| 2 | Between 36-40 \| N/A \| 1 subject | 1M | N/A | African American |
| 2 | 48.71 \| 6.92 \| 7 subjects | 3F/4M | 42.85%/57.14%/ | Arab/Middle Eastern |
| 2 | 29.28 \| 6.92 \| 7 subjects | 4F/3M | 57.14%/42.85%/ | Asian |
| 2 | 43.74 \| 17.94 \| 55 subjects | 32F/23M | 58.18%/41.81% | Caucasian |
| 2 | 35.33 \| 9.266 \| 6 subjects | 3F/3M | 50%/50% | Hispanic/Latino |
| **Totals** | 76 controls | 55% F/ 44% M | 52% F/ 47% M | 1.32% African American/ 9.21% Arab / 9.21% Asian / 72.37% Caucasian / 7.89% Hispanic |

Each patient participated in two SPECT brain scans, acquired during rest and while performing a Conners Continuous Performance Test (Conners Continuous Performance Test, CCPT-II, Multi-Health Systems, Toronto, Ontario; Conners, 2000) across 14 clinical imaging sites. SPECT scans were acquired using Picker (Philips) Prism XP 3000 triple-headed gamma cameras (Picker Int. Inc., Ohio Nuclear Medicine Division, Bedford Hills, OH, USA) or InterMedical MultiCam 3000eco triple-headed gamma cameras (Intermedical Medizintechnik GmbH, Lubbecke, D-32312, Germany) with low energy high resolution fan beam collimators.

For each procedure, an age- and weight-appropriate dose of 99mTc–hexamethylpropyleneamine oxime (HMPAO) was administered intravenously at rest. For the rest scans, patients were injected while they sat in a dimly lit room with their eyes open. Patients were scanned for approximately 30 minutes after injection. Data acquisition yielded 120 images per scan with each image separated by three degrees, spanning 360 degrees. A low pass filter was applied with a high cutoff and Chang attenuation correction performed (L.-T. Chang, 1978; W. Chang et al., 1984). The resulting reconstructed image matrices were 128×128×78 with voxel sizes of 2.5mm^3^.

For voxel-based analyses, images were aligned to the Montreal Neurological Institute (MNI) space with the Advanced Normalization Tools (ANTs version 2.2.0; Avants et al., 2011); RRID:SCR_004757) using a SPECT template, resulting in an image matrix size of 79×96×68 with isotropic voxel sizes of 2.0mm^3^. SPECT images were scaled to the within-scan maximum voxel and noise outside of the brain was removed using a 50% of maximum threshold, prior to registration. After the thresholded images were aligned to the MNI space and the transformation applied to the un-thresholded images, a brain mask derived from the MNI 152 (Fonov et al., 2011) template was used to remove noise outside the brain from the un-thresholded images for use in the statistical models. Registered SPECT scans were visually checked for the absence of severe functional abnormalities or artifacts and proper registration to the MNI space.

### ICA Processing

Preprocessed SPECT data were analyzed via spatially constrained ICA using the 68 component NeuroMark ICA SPECT template (Harikumar et al., 2026; *in prep*) through the Group ICA of fMRI Toolbox (GIFT; http://trendscenter.org/software/gift; RRID:SCR_001953). Analysis was performed using the GSU ARCTIC high performance computing cluster (https://arctic.gsu.edu/; see acknowledgment section for more information). Spatial montage maps and initial statistical analyses were run with MATLAB R2020b (Version 9.9.0.2037887). 2,819 subjects (healthy controls and depressed patients combined) were reduced to 68 principal components after performing a principal component analysis (PCA) analysis, and an sc-ICA was subsequently performed using the MOO-ICAR algorithm (Du et al., 2019, 2020).

### Statistical Analyses

Following this initial analysis, a correlation plot was created to visualize clinical correlates between the transdiagnostic patient groups and Amen Brief Symptom Checklist and General Symptom Checklists (BSC; GSC; https://ancronmedical.com/wp-content/uploads/2015/04/Amen-Adult-General-Symptom-Checklist.pdf). Next, pairwise correlations between the loading parameters for the SPECT components were analyzed for within and between group differences in Python 3.15 using the ttest_ind package from SciPy (https://docs.scipy.org/doc/scipy/reference/generated/scipy.stats.ttest_ind.html). Regressions corrected for multiple comparisons using the false discovery rate (FDR; Benjamini & Hochberg, 1995) on loading parameter values between both groups were performed to identify group differences.

We then regressed these results against typical demographic variables such as age, sex, clinic location, and three factors associated with common depression symptoms, such as worry/rumination, moodiness, and social disinterest, with primary regression analyses, bar plots, and figures generated in Python 3.15 with an FDR correction utilized using the fdr_bh function in the statsmodels package (https://www.statsmodels.org/dev/about.html). A regression was run using the following framework, and results were FDR corrected for multiple comparisons, with the equation utilized below:

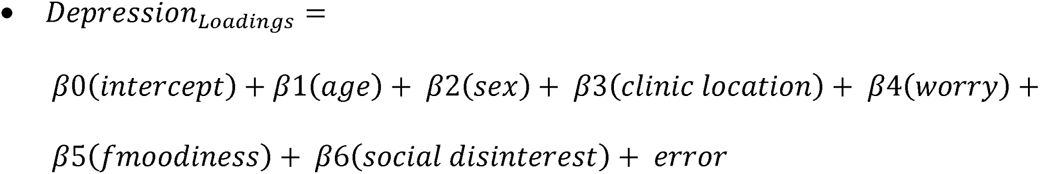

## RESULTS

### 1. Clinical Symptom Correlation Matrix Results

First, we investigated clinical symptom correlations across the six groups, and between BSC and GSC select items (Figure 4). We selected certain items of interest that relate symptom wise across all six group profiles due to previous evidence of cognitive/executive functioning deficits in these groups (Alves et al., 2014; Qi et al., 2020a; Schwarz et al., 2020). Unsurprisingly, moderate correlations were noted regarding the BSC_45 (“Tending to predict fear”) and GSC_18 (“Experiencing panic attacks”) items and anxiety disorder. Additional moderate correlates were found between BSC_78 (“Loss of motivation”) and GSC_99 (“Thoughts of speech changes”), BSC_59 (“Lowered interest”) and BSC_78 (“Loss of motivation”), Interestingly, GSC_99 (“Thoughts of speech changes”) was moderately correlated with GSC_8 (“Feelings of worthlessness”; *r = 0.46).* Additionally, fear-based items (fear of dying vs. predict fear) were moderately correlated; *r = 0.46*.

**Figure 4.**
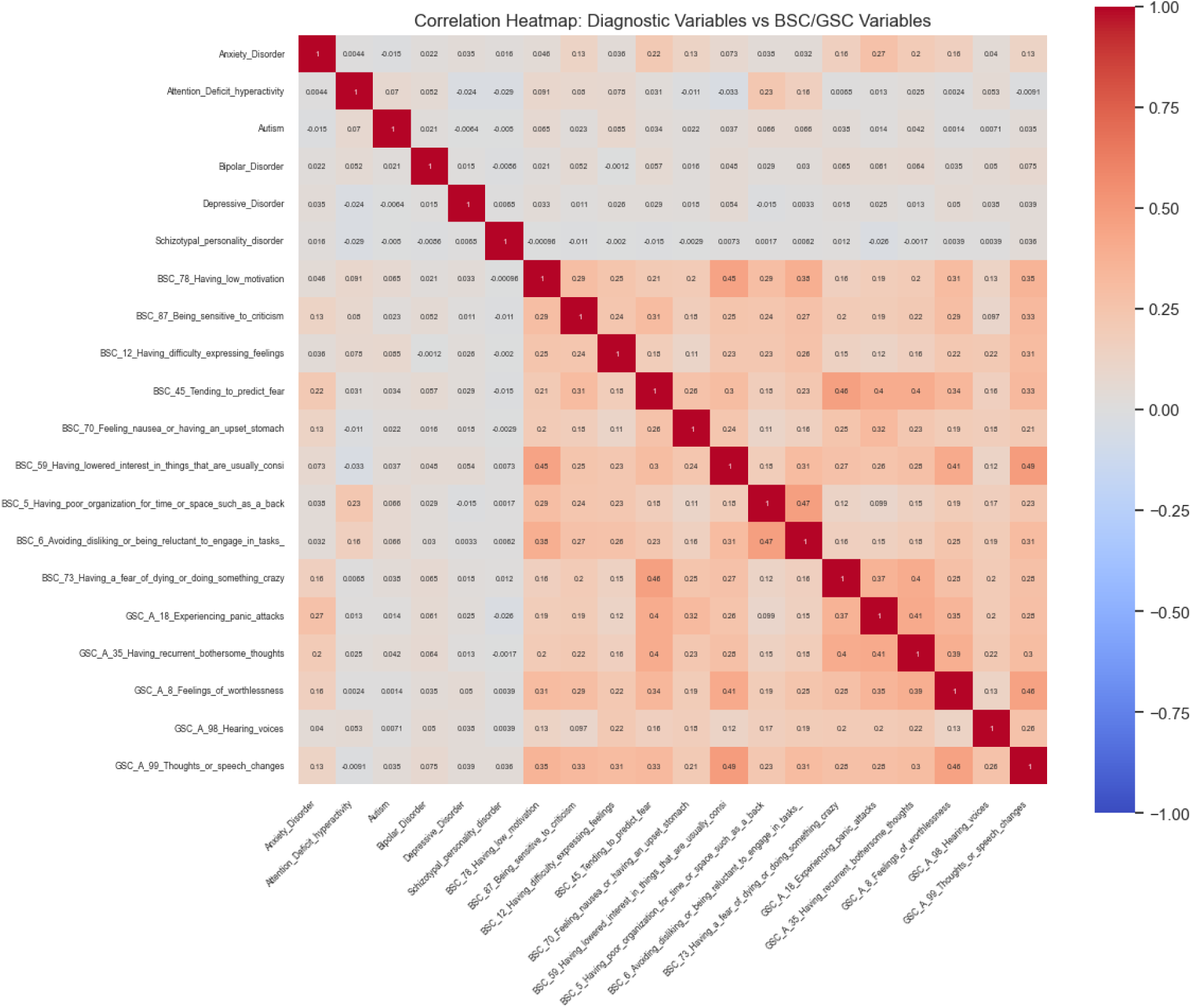
Correlation plot showing anxiety/depression oriented BSC/GSC variables correlated against the six diagnostic groups. Unsurprisingly, results showed increasingly moderate correlations for items related to panic attacks, fear, and upset stomach in individuals with anxiety. Consequently, other items related to decreased interest, fear of dying, and reluctancy to complete tasks were more correlated with each other.

### 2. Loading Parameter Group Differences

We examined the loading parameter patterns between healthy controls vs. patients (Figure 5a) with positive (HC > DEP) and negative (DEP > HC) components plotted. We observed that across the various NeuroMark domains and subdomains that 31 components (Figure 5b) as displayed below were significant at both the *q< 0.05* and *q < 0.01* thresholds. A variety of domains/subdomains across the cerebellar, higher cognition-frontal (HC-FR), higher cognition – temporoparietal (HC-TP), higher cognition – insular temporal (HC-IT), subcortical – extended thalamus (SC-ET), subcortical – extended hippocampus (SC-EH), subcortical – basal ganglia (SC-BG), subcortical – extended thalamus (SC-ET), sensorimotor (SM), triple network – central executive (TN-CE), triple network – default mode (TN-DM), triple network – salience (TN-SA), visual-occipital (VI-OC) and visual – occipitotemporal (VI-OT) per the NeuroMark 2.2 labeling conventions (Jensen et al., 2024) were represented in the results. Controls had increased loadings broadly represented across the cerebellum, HC-FR, HC-IT, HC-TP, TN-DM, TN-CE, and VI-OC domains and subdomains. By contrast, patients had increased loadings selectively in certain domains/subdomains across cerebellar, SC-BG, SC-EH, SC-ET, TN-DM, and VI-OC components. In particular, patients had less within domain/subdomain component representation compared to healthy controls, which may be reflective of segregated network dysfunction and disruptions related to certain cognitive and executive networks.

**Figure 5a.**
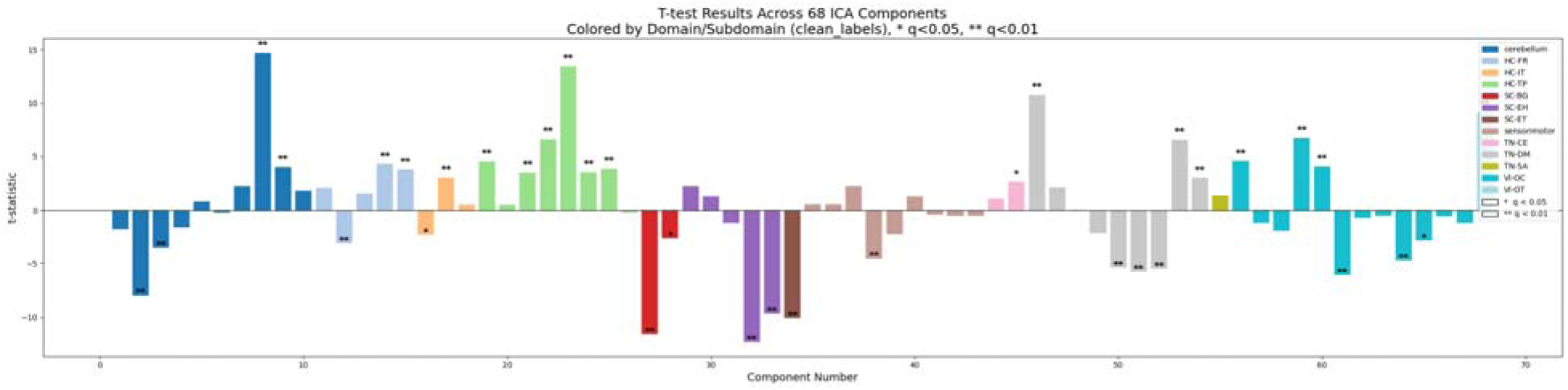
Group differences between healthy-patient loadings showed 31 broader domain/subdomain significant group differences for components represented across 13 NeuroMark 2.2 domains/subdomains. Patients had more segregated component loadings (e.g. fewer loadings per domain/subdomain) compared to healthy controls in cerebellar, SC-BG, SC-EH, SC-ET, TN-DM, and VI-OC domains/subdomains, which could be indicative of segregated network dysfunction compared to controls.

**Figure 5b.**
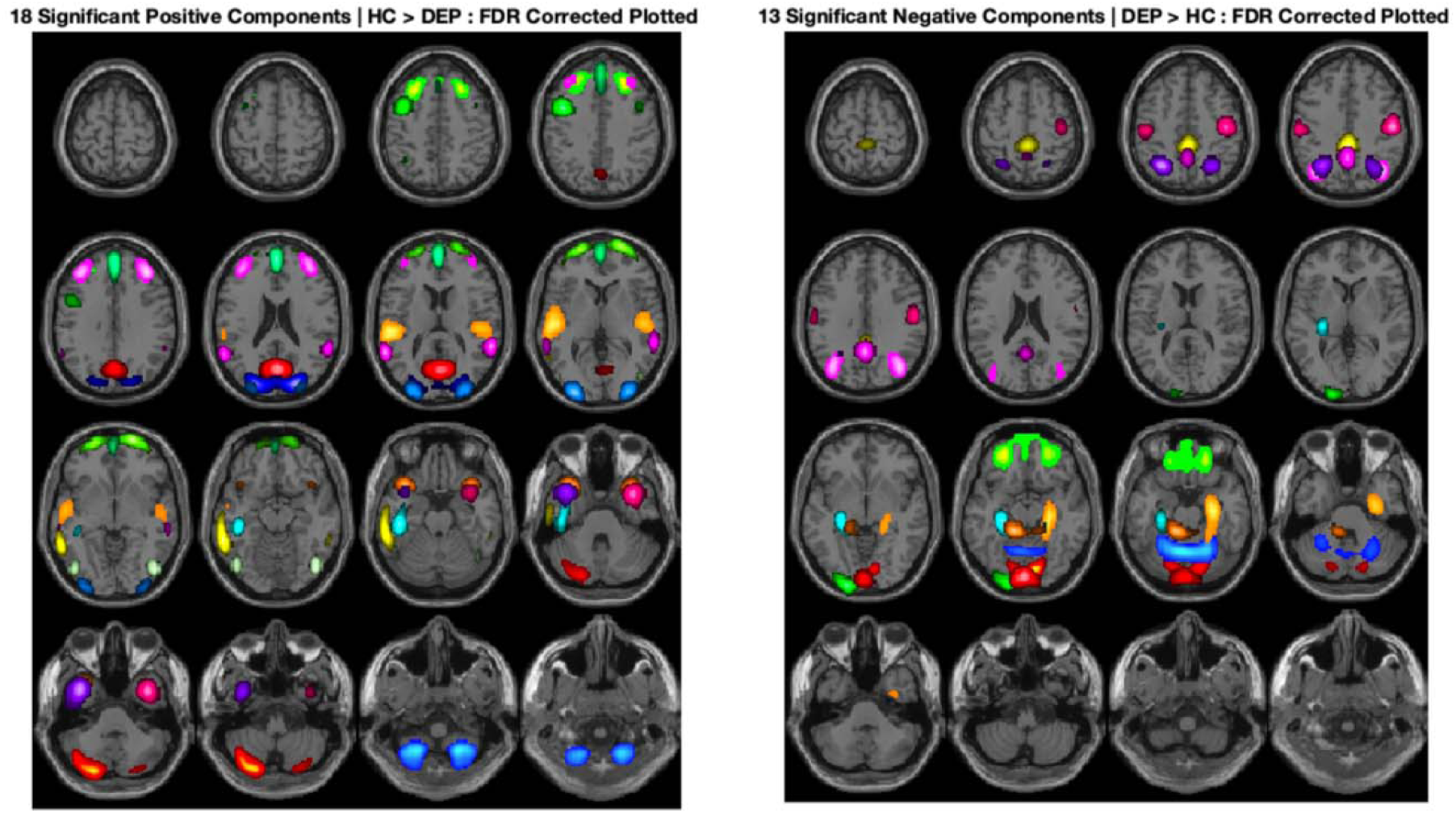
Spatial montage map display demonstrating the 31 components (18 HC > DEP, 13 DEP > HC) that remaining significant after q <0.05. In particular, patients had more segregated component loading (e.g. fewer loadings per domain/subdomain) compared to healthy controls in cerebellar, HC-FR, HC-TP, HC-IT, SC-BG, SC-EH, SC-ET, sensorimotor, TN-DM, TN-SA, and VI-OC domains/subdomains), which could be indicative of segregated network dysfunction compared to controls.

### 3. Clinical Outcomes: Measuring the effects of Age, Sex, Clinical Site, and Factor Analysis Variables on Depression Loadings

#### 3a. Worry/Rumination

We first examined worry/rumination as the first factor (Figures 6a-6c) within depression symptoms, given that this is a hallmark symptom of the disease (McLaughlin et al., 2007). Overall, we noted that after performing FDR correction, cerebellar, SC-BG, SC-EH, and sensorimotor components were positively associated with worry. By contrast, HC-FR, HC-TP, and sensorimotor components were negatively associated with worry/rumination behaviors.

**Figures 6a-6b.**
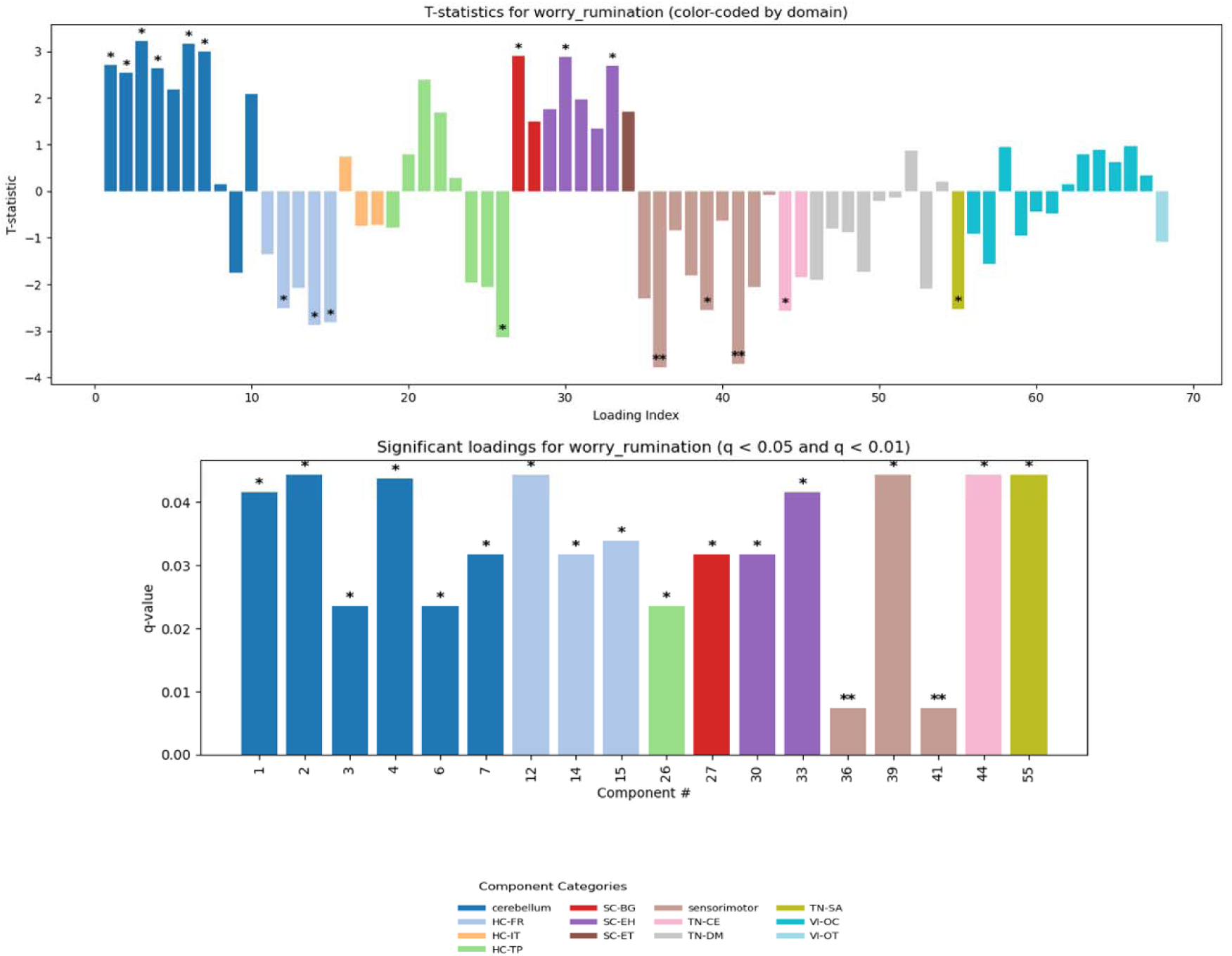
Figure 6a shows significant components plotted with FDR correction employed at q-< 0.05 and q < 0.01. Figure 6b shows the specific components that survived FDR correction more closely. Positive correlations were associated with cerebellum, SC-BG, SC-EH, and sensorimotor domains/subdomains. Negative correlations were associated with the HC-FR, HC-TP, and sensorimotor components.

**Figure 6c.**
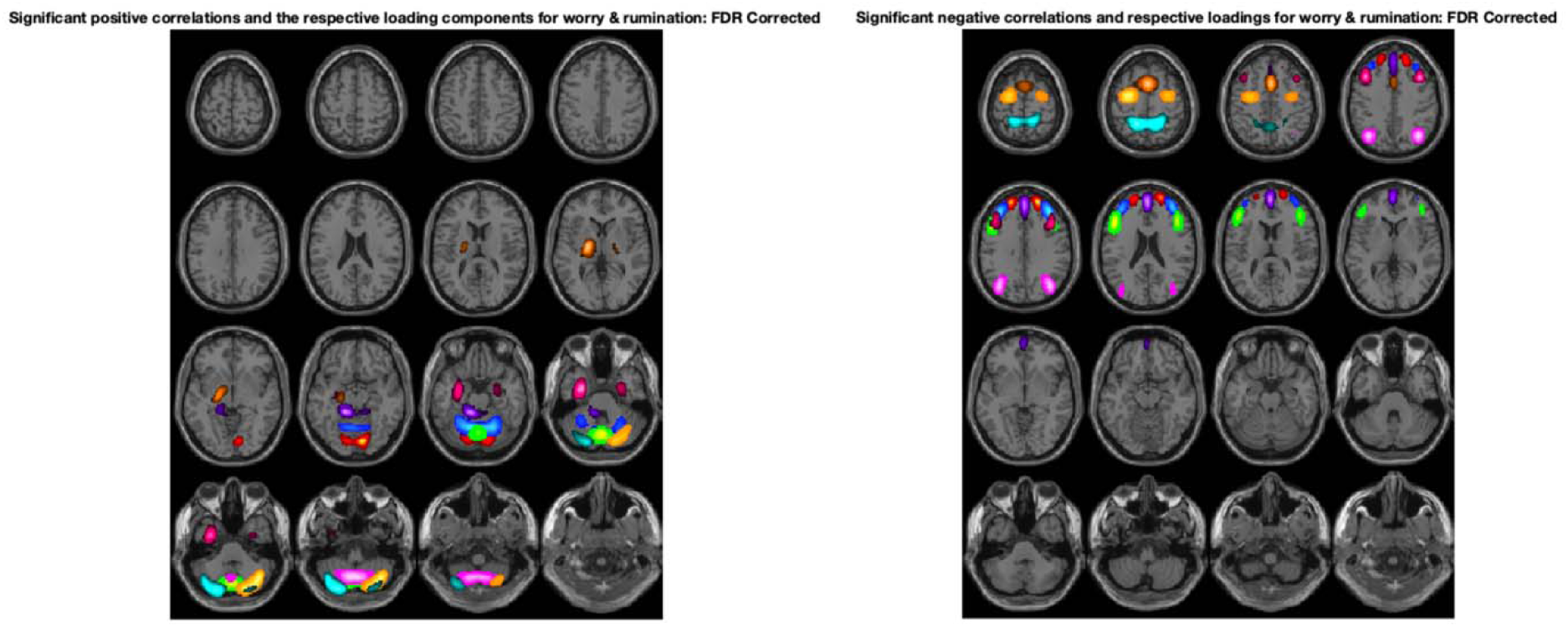
shows a spatial montage map of the significant components plotted in Figure 6b, demonstrating activity in the cerebellar and subcortical networks (i.e. cerebellum, SC-BG, SC-EH networks) positively with worry; conversely, activity in the HC-FR, HC-TP, and sensorimotor networks were negatively correlated with worry and rumination symptoms.

#### 3b. Moodiness

Next, we looked at moodiness which is a classic symptom in depression (Figures 7a-7b). Overall, t-statistic plots show that broadly, components were evenly spread across all 11 domains/subdomains. No significant components associated with moodiness survived FDR correction.

**Figures 7a-7b.**
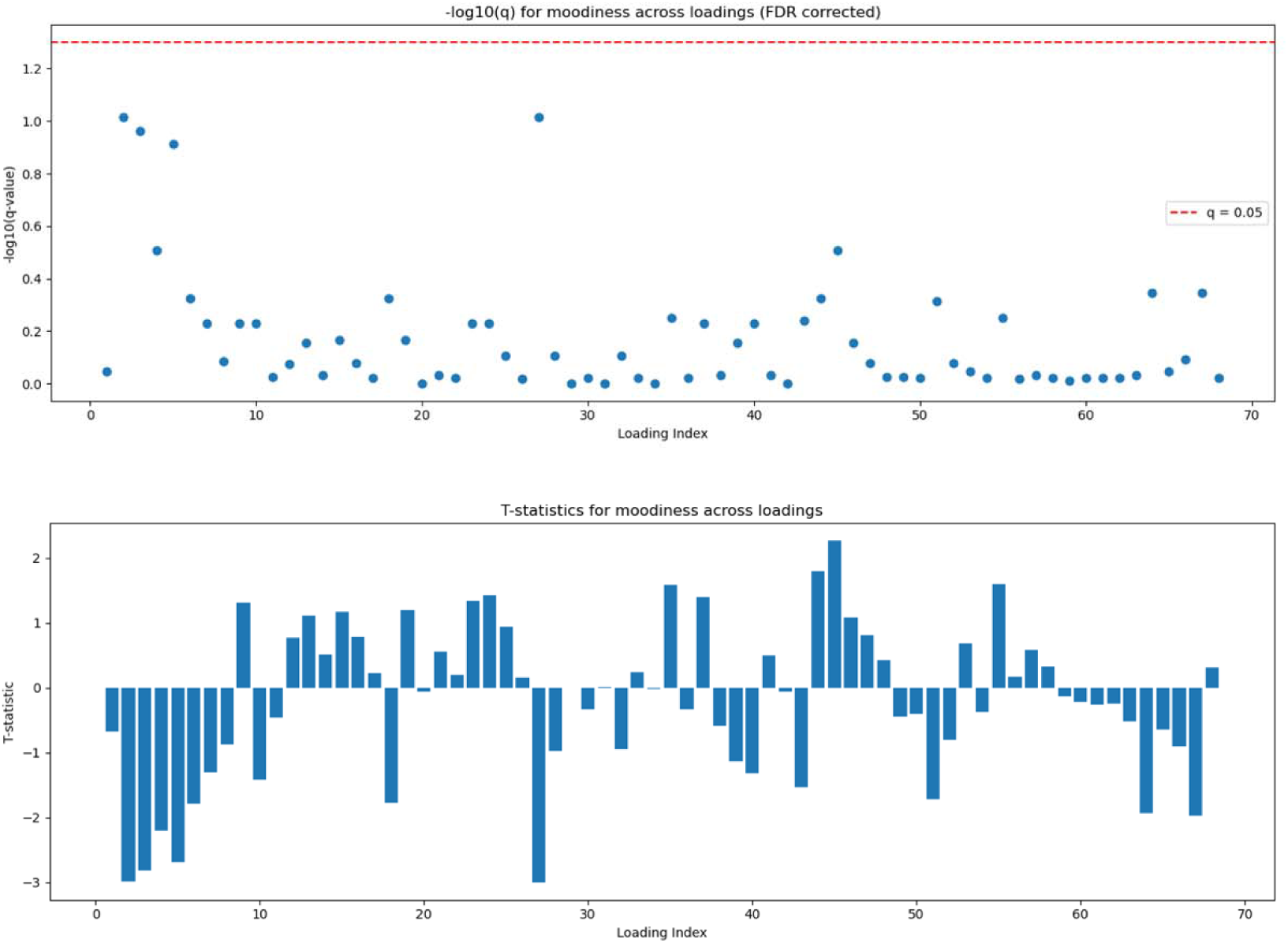
Figure 7a(top) shows scatter spread of -log10(q) values for the regression to demonstrate the spread of the data. As per Figure 7a, no component results survived FDR correction.

#### 3c. Social Disinterest

Finally, social disinterest was considered (Figures 8a-8c) given the fact that anhedonia and lack of motivation to pursue interests is a critical piece in disorders such as schizophrenia, depression, and autism (Barkus & Badcock, 2019). We noted that cerebellar, HC-TP, TN-DM, TN-SA, and VI-OC components were the most significant components associated with social disinterest compared to others, with TN-SA being more prominently expressed in based on the t-statistic patterns. Component 55 (TN-SA) was significantly correlated with social disinterest symptoms.

**Figures 8a-8b.**
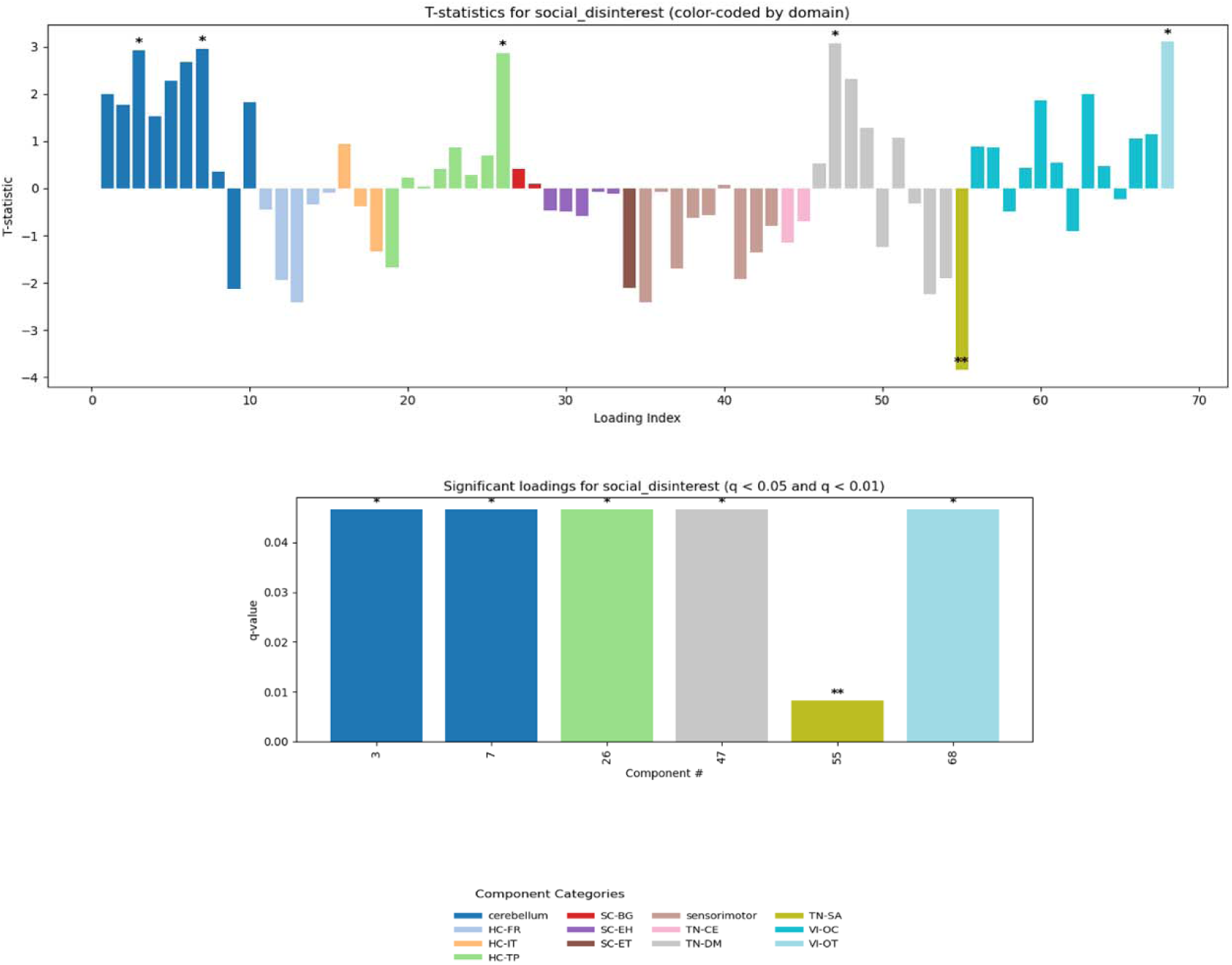
Figure 8a provides significant components of interest, including cerebellar, HC-TP, TN-DM, TN-SA, and VI-OC components. Figure 8b shows the components which survived FDR correction more closely. Positive correlations were associated with cerebellar, HC-TP, TN-DM, and VI-OT domains and subdomains. Notably, Component 55 (TN-SA) had a strong negative association with social disinterest symptoms in patients.

**Figure 8c.**
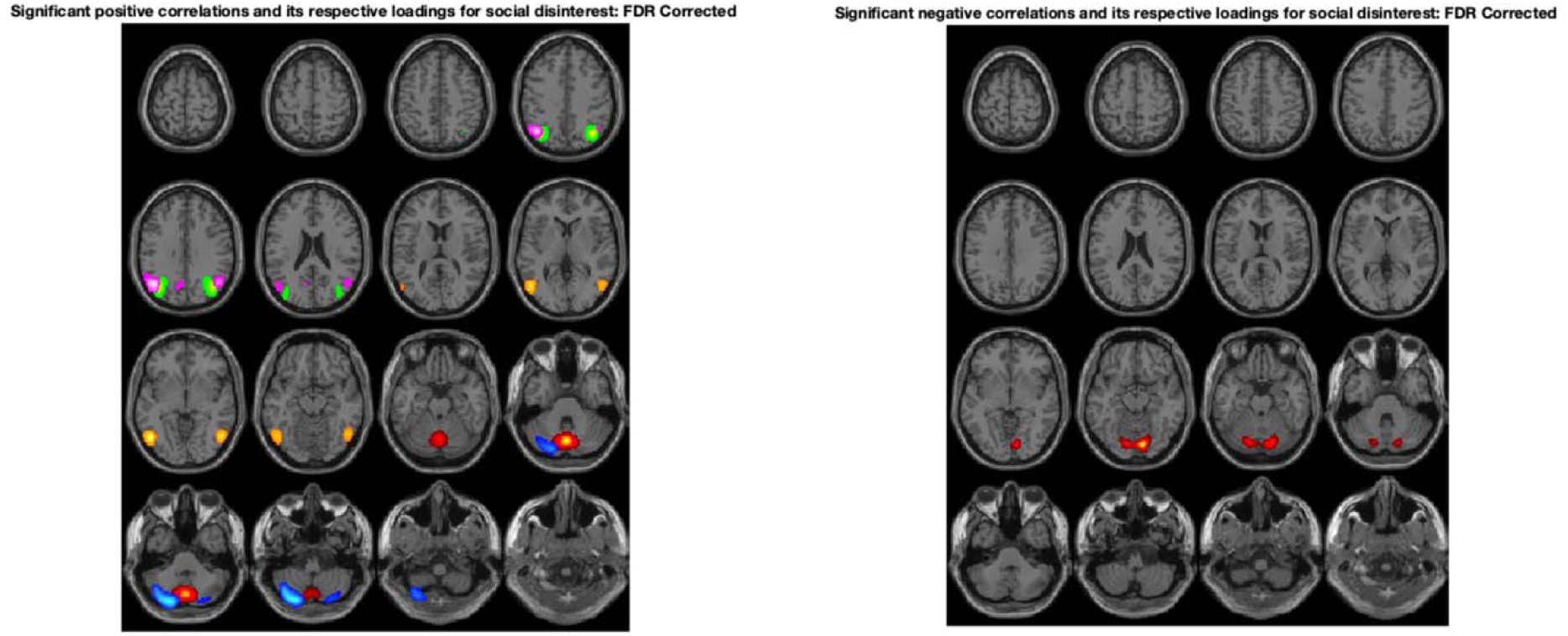
Figure 8c demonstrates the components plotted in Figure 8a; notably, components across the cerebellar, HC-TP, TN-DM, TN-SA, and VI-OC domains/subdomains were associated with social disinterest. Positive correlations with social disinterest and depression loadings were associated with HC-TP, TN-DM, cerebellar, and VI-OT domains and subdomains. Conversely, negative correlations were associated with the TN-SA subdomain.

## DISCUSSION

These preliminary findings demonstrate the utility of a combined SPECT + NeuroMark template based sc-ICA approach for increased predictive power. We identified unique patterns of loading parameter expression broadly between patients and controls across cerebellar, frontal, sensorimotor, and visual domains/subdomains. This suggests interesting segregation of domains/subdomain differences particularly in patients compared to controls.

These results are unsurprising, given that previous literature has shown a wealth of evidence related to specific networks associated with anxiety and depressive symptoms (Abdoli et al., 2022; J. Li et al., 2018; Z. Li et al., 2021; Liu, 2025; McLaughlin et al., 2007; Qi et al., 2020a; Richieri et al., 2015; Schwarz et al., 2020; Sridhar et al., 2026; Sylvester et al., 2012). Our study represents the first transdiagnostic SPECT study, to our knowledge, that separates patient vs. control trends across cognitive and executive function related networks. Furthermore, we showed domain/subdomain specific sensitivity as noted in the loading parameter analysis, with controls having more widespread loading expression across a variety of HC, subcortical, and triple network regions. Patients, however, showed a more restricted, segregated pattern which is consistent with previous fMRI literature related to default mode network and frontal network dysconnectivity (Sylvester et al., 2012). Additionally, given that SPECT is a measure of rCBF, we noted that the SPECT data was able to capture a variety of group differences across loading parameters. These have been seen in prior schizophrenia studies (Harikumar et al., 2026, in prep; Harikumar et al., 2025) and highlights increased clinical utility of SPECT for understanding psychiatric disorders.

We also identified specific components associated with each of the three factor analyses performed for worry/rumination, moodiness, and social disinterest with most results identifying cerebellar, subcortical, triple network, and visual components, which are unsurprising. There are many possibilities as to why these components were significant. Prior transdiagnostic neuroscience child/adolescent studies have shown deficiencies in ADHD, depression, and anxiety related to decision making and emotion processing (Sonuga Barke et al., 2016). Given that individuals with these disorders have impaired reinforcement learning and emotional processing abilities, the components we identified could be important in identifying clinical impairments (e.g. over recruitment of certain brain regions, impairments in higher cognition related areas) which need further examination. Prior studies have also identified sensorimotor differences in ADHD patients vs. healthy individuals (Duerden et al., 2012), particularly altered cortical thickness and thinning frontal lobe structured in ADHD, which could be contributing to the group differences we noted in our study. Additionally, atypical functioning of cortico-cerebellar, corticostriatal systems could play a part in the additional identification of cerebellar/subcortical components we noticed in our findings, as seen in previous fMRI studies (Valera et al., 2010), particularly in ADHD and depressed populations (Felger et al., 2016). We also identified aberrant cortico-striatal patterns in prior SPECT studies, especially in schizophrenia populations (Harikumar et al., 2025) which linked aberrant patterns to auditory hallucination symptoms. These existing atypical and aberrant patterns of dysconnectivity could be driving the group differences observed here. However, more SPECT studies are needed to fully understand and appreciate clinical differences between transdiagnostic populations across disorders such as autism, depression, bipolar disorder, and psychotic disorders.

One limitation in this study was the imbalanced patient/control sample sizes. However, we were able to leverage a sample size of over 2,700 SPECT patients which is a significant strength. Additionally, this study was able to find correlations between clinical symptoms and across functional networks, replicating prior trends from studies. Future studies should utilize a larger sample size, as well as more robust clinical measures and group comparisons for further review.

## CONCLUSIONS

Taken together, these results represent the first transdiagnostic SPECT study of its kind utilizing a novel NeuroMark SPECT template approach. While many results followed expected patterns, the loading parameter analyses coupled with our correlation results suggest specific domain/subdomain network segregation for depressed patients along with expected patterns of cortical-subcortical and higher cognition/executive network dysfunction across certain groups in our study.

## Data Availability

Due to the sensitive nature of this clinical data, the SPECT data has not been posted publicly but the anonymized demographic and other clinical data used in this study is freely available for research by request to Dr. Keator. The code in MATLAB and other associated files are available on Github: (https://github.com/trendscenter/gift-bids/tree/main/misc/spect/proj/march2024).

https://trendscenter.org/data/

## Acknowledgments

We acknowledge the use of Advanced Research Computing Technology and Innovation Core (ARCTIC) resources at Georgia State University’s Research Solutions made available by the National Science Foundation Major Research Instrumentation (MRI) grant number CNS-1920024.

## Author Contributions

AH was responsible for primarily preparing, drafting, and writing the manuscript, along with conducting the analyses and generation of all figures, tables, and references.

BTB, DA and DK were responsible for providing feedback, access to SPECT data, funding, edits of the manuscript, and reviewed all figures, tables, and references.

VDC were responsible for conceptualization, supervision, guidance, funding, and primary edits for the manuscript and template creation process.

All authors reviewed the manuscript.

## Statement of Informed Consent/Approval

Integ Review IRB, Protocol: 004-Amen Clinics Inc., Determination: Exempt Category 4. Approval Date: 9/19/2014. Study Title: Retrospective Review of Clinical Cases in a Brain SPECT Imaging Database. Investigator: Daniel Amen, M.D. Integ Review IRB was acquired by Advarra IRB.

## Declaration of Generative AI and AI Assisted Technologies in the Writing Process

During the preparation of this work the primary author used Grammarly, Microsoft Copilot, and ChatGPT to check content for flow and clarity, verify simple summation totals for tables, create and streamline MATLAB code for efficiency, and proofread the work for grammar and spelling errors. After using this tool/service, the primary author reviewed and edited the content as needed and take(s) full responsibility for the content of the published article.

